# Genomic insights into early SARS-CoV-2 strains isolated in Reunion Island

**DOI:** 10.1101/2021.01.21.21249623

**Authors:** David A Wilkinson, Camille Lebarbenchon, Célestine Atyame, Sarah Hafsia, Marie-Christine Jaffar-Bandjee, Luce Yemadje-Menudier, Sébastien Tanaka, Olivier Meilhac, Patrick Mavingui

## Abstract

The relative isolation of many island communities provides some protection from the COVID-19 pandemic, as imported cases can be limited and traced effectively. Until recently, this was true for the population of the French overseas department, Reunion Island, where only limited numbers of autochthonous cases were observed prior to August 2020. Since the report of the first case of COVID-19, contact tracing has been carried out for each new case identified in Reunion Island to identify transmission and clusters. To contribute to the public health response and understand the diffusion of SARS-Cov-2 strains in Reunion Island, we established in-house genome sequencing capability in Reunion using Oxford nanopore technology (MinION) as an inexpensive option for genomic typing of SARS-CoV-2 lineages on the island, and cross-validated typing results between viral isolation methods and different sequencing technologies. The results of our work during the early phase of the epidemics are presented herein.

**Article Summary Line:** The COVID-19 pandemic has had an unprecedented impact on the global community. Here we provide epidemiological and genomic details of the early stages of the pandemic on Reunion Island.

## Introduction

Severe acute respiratory syndrome coronavirus 2 (SARS-CoV-2) was first identified as the causative agent of coronavirus disease 2019 (COVID-19) in China in December 2019 after its emergence from wild animal populations[1]. Since, it has infected more than 45,000,000 individuals with 1.2 million deaths worldwide (at the time of writing). Its transmission has been rapid and widespread despite global intervention measures (such as quarantine and travel restrictions) due largely to the fact that viral transmission can occur from asymptomatic individuals and pre-symptomatic individuals[2]. This coupled with its relatively high case fatality rate[3] means that SARS-CoV-2 remains a public health emergency of unprecedented scale.

The response to this pandemic has also been of an unprecedented scale. Governments have restructured healthcare systems to accommodate increased testing capacity for improved contact tracing, and genomics has played an important role in improving our understanding of COVID-19 transmission pathways and outbreaks[4,5]. As the number of acquired mutations in the SARS-CoV-2 genome has been relatively low over the course of the pandemic, whole genome sequencing has been essential to allow accurate strain typing, which in turn allows cases to be associated based on genetic similarity of the infecting strains. One study from Australia estimated that the use of genomic typing in conjunction with classical epidemiological tracing improved the case assignment rate by as much as 16%[6]. Next generation sequencing has therefore been of great importance during the SARS-CoV-2 pandemic and emerging technologies such as Oxford Nanopore sequencing have made genomic approaches more accessible: To date, there have been more than 100,000 SARS-CoV-2 genomes deposited in the Global Initiative on Sharing All Influenza Data (GISAID) database[7].

Reunion Island is an overseas department of France situated in the Indian Ocean and is largely subject to the same regulations concerning COVID-19-related practices as mainland France and the other French overseas departments. Reunion Island has a population of approximately 850,000 people who live mainly in coastal regions. The population is relatively young, with people 65 years or over representing 8.1 % fewer of the population than in mainland France (2020 institute of national statistics data, www.insee.fr/fr/statistiques). The first case of COVID-19 was reported in Reunion on 11th March 2020, four days before French regulations imposed strict lockdown measures which significantly reduced mobility across the country[8]. Prior to this in February, all direct commercial flights between Reunion and China had been suspended. In addition, lockdown measures provided effective protection for the Reunion population, meaning that numbers of cases remained low and were mainly imported as local inhabitants returned from areas of active SARS-CoV-2 circulation. Importantly, during this time most inbound flights were canceled, returning passengers were obliged to undergo a 14-day confinement period upon arrival, and all returning individuals were closely monitored by regional health authorities. Limited local virus circulation occurred in Reunion at this time, with approximately 70 % imported cases and 30 % autochthonous cases (defined as most likely local transmission with no history of travel), with a limited severity [9]. Subsequently, Reunion avoided the first pandemic wave that swept through Europe and placed the European population at the epicenter of the pandemic from March to April 2020. A timeline of reported cases and important events in the epidemiology of COVID-19 on Reunion Island is presented in Figure 1. Here we isolated and provided genomic data of SARS-CoV-2 strains circulating during the early phase of the epidemics on Reunion Island.

**Figure 1:**
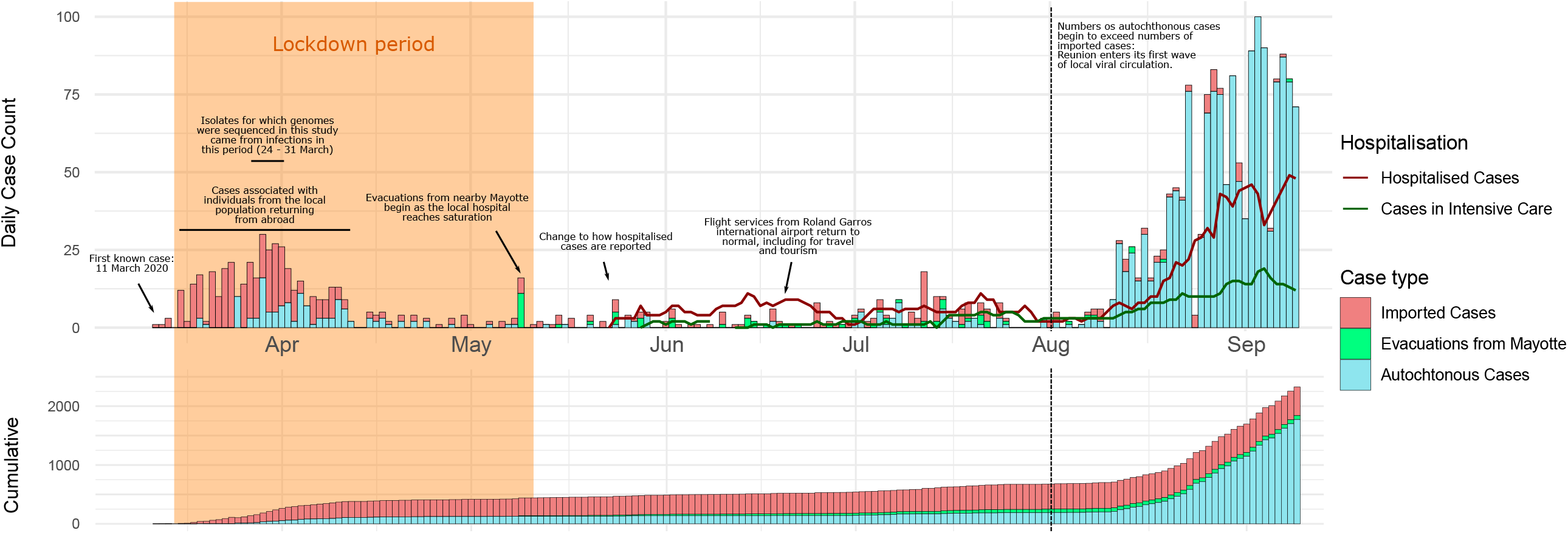
Epidemiological timeline of COVID-19 cases in Reunion Island corresponding to the first six months since the first identified cases (11th March 2020 – 11 September 2020).

## Methodology

### Patients and samples

Nasopharyngeal specimens were recovered either from patients admitted to CHU Reunion or from outpatients, all with laboratory confirmed COVID-19. Before use, we ascertained the presence of viral genomic ARN in each specimen swab by real time RT-qPCR (described below). Samples were handled and stored at −80°C in a biosecurity level three virology laboratory. Consent was obtained from each patient, classified as non-opposition, at the time of entry to the hospital.

### RNA Extraction and RT-qPCR

Nucleic acids were extracted using the QIAamp Viral RNA mini kit (QIAGEN, Valencia, California, USA) and eluted in 60 μL of AVE buffer. Reverse transcription was performed on 10 μL of RNA using the ProtoScript II Reverse Transcriptase and Random Primer 6 (New England BioLabs, Ipswich, MA, USA) as previously described[10]. cDNAs were tested for the presence of the N and Nsp14 genes using the PCR protocol published by The University of Hong Kong (https://www.who.int/publications/m/item/molecular-assays-to-diagnose-covid-19-summary-table-of-available-protocols). PCR were performed with the QuantiNova Probe RT-PCR Kit (QIAGEN, Valencia, California, USA) and 5 µL of cDNA in a CFX96 Touch™ Real-Time PCR Detection System (Bio-Rad, Hercules, CA, USA).

### Viral isolation and production

SARS-CoV-2 strains were isolated in the African green monkey kidney cell line, Vero E6. Briefly, cells were cultured in a 25 cm^2^ flask containing 5 ml of Eagle medium (MEM: Gibco/Invitrogen, Carlsbad, CA, USA) supplemented with 5% heat-inactivated fetal bovine serum (FBS), 2 mmol/L L-Glutamine, 1 mmol/L sodium pyruvate, 100 U/mL of penicillin, 100 µg/mL of streptomycin and 0.5 µg/mL of Amphotericin B (PAN Biotech, Aidenbach, Germany), at 37°C under a 5% CO_2_ atmosphere. When cell density reached approximately 80%, cells were rinsed once with MEM medium supplemented with 2% FBS, then 50 to 100 µl of SARS-CoV-2-positive swab specimen was mixed with 2 mL MEM 2% FBS and layered on the Vero cell monolayer. After 2 h of incubation the inoculum was removed, cells were rinsed once and 6 mL of fresh MEM 2% FBS medium was added. Flasks were incubated at 37°C in a 5% CO_2_ atmosphere. Cultured were checked daily. Cell lines demonstrated a cytopathic effect 3 to 5 days post-infection. Supernatants of positive cultures were recovered, centrifuged at 400 X g for 5 min at 4°C, then aliquoted and stored at −80°C until used.

For viral production, a total of 100 µl supernatant of positive culture was used to infect a monolayer of Vero cells grown in 75 cm2 flask as above. After three days, cytopathic effect was visible and the supernatant was recovered, cleared by centrifugation, aliquoted and stored at − 80°C.

### Viral titration

Titration was performed by plaque-forming assay. Vero cells (cultured as above) were seeded in 6-well culture plates. Tenfold dilutions of SARS-CoV-2 solution were made in MEM 2% FBS culture medium and 500 µl of each dilution was added to the cells in duplicate. After an incubation of 2 h at 37°C under a 5% CO_2_ atmosphere, unabsorbed viral particles were removed and 2 ml of culture medium supplemented with 0,8% carboxymethylcellulose (CMC; Sigma-Aldrich, Saint-Quentin-Fallavier, France) was added to each well and incubation was extended for 5 days. The CMC overlay was carefully removed, and the cells were fixed with 2 mL of 3.7 % paraformaldehyde in PBS. The plates were incubated at 37°C for 20 min. The cells were then rinsed once with PBS and stained with 0.5% crystal violet in 20% ethanol. Plaques were visually counted and expressed as plaque-forming units per mL (PFU/mL).

### Nanopore (MinION) Sequencing and Analysis

Genomes were sequenced from viruses originally infecting four independent patients. Two samples (RUN-PIMIT1 and RUN-PIMIT2) were from couple of travelers returning from France mainland. Sample RUN-PIMIT8 was obtained from a tourist originating from Greece whereas sample RUN-PIMIT20 was quoted as real autochthonous case with unidentified contact with an imported case. We combined different sequencing technologies to cross-validate the sequences generated by different protocols. We also re-sequenced the same isolates from different passage histories to examine whether cell-line isolation may result in the accumulation of mutations.

In total, ten SARS-COV2 genomes were sequenced on a single Oxford Nanopore Technologies FLO-MIN106D (R9.4.1) flowcell using the Artic Network’s overlapping amplicon protocol[11,12]. Following first-strand cDNA synthesis (as above), all sample preparation methods followed the nCoV-2019 sequencing protocol version 2 (https://dx.doi.org/10.17504/protocols.io.bdp7i5rn). Briefly, genomic material was amplified in two independent PCR reactions using version 3 primer pools obtained from of the Artic network website (https://github.com/artic-network/artic-ncov2019). PCR products were pooled, dA-tailed using the NEBNext® Ultra™ II End Repair/dA-Tailing Module then barcoded using the Nanopore Native Barcoding Expansion kit (EXP-NBD104). Barcoded amplicons were then purified using a 0.4x volume of AMPure-XP SPRI beads, washing the beads with an excess volume of Nanopore’s small fragment buffer (SFB) to ensure that un-ligated barcode molecules were removed. Purified amplicons were then pooled in equimolar proportions before adapter ligation and sequencing.

The sequencing run was left for 24 hours and stopped when predicted coverage exceeded 1000-fold for each genome. Basecalling and demultiplexing were performed using guppy (v4.0.11). Basecalling used default parameters in accurate mode, and demultiplexing was performed using the “--require_barcodes_both_ends” parameter to minimize sample crosstalk. Reads were then assembled using Medaka v1.0.3 (https://nanoporetech.github.io/medaka/) as part of the Artic network bioinformatics standard operating procedure (https://artic.network/ncov-2019/ncov2019-bioinformatics-sop.html), subsampling amplicon coverage to 400x and using genome accession MN908947.3 for read mapping. Geneious v9.1.8[13] was used to inspect and curate mapped sequence data. Consensus base-calling required a minimum of 30-fold coverage.

### Illumina Sequencing and Analysis

Four of the SARS-CoV-2 samples (2 isolates and 2 swabs) that were sequenced using the ONT amplicon protocol were additionally sequenced using an Illumina shotgun method from unamplified cDNA in order to complete genomic loci that were poorly amplified by the tiling PCR and to allow cross validation of the ONT sequencing methodology against the Illumina gold standard.

Briefly, sequencing libraries were generated from 10 ng of cDNA using the Celero™ PCR Workflow with Enzymatic fragmentation (DNA-Seq) following the manufacturer’s instructions. Sequencing was performed on the MiSeq platform, with 1 * 170 bp single end reads. Demultiplexed sequences were provided by the sequencing company (Biofidal, Lyon, France). Sequences were quality trimmed and adapters removed using Trimmomatic v0.39. Trimmed reads were mapped to reference sequence MN908947.3 using minimap2 v.2.17-r941[14]. Geneious v9.1.8[13] was used to inspect and curate mapped sequence data and consensus sequences.

### Genome typing

Assembled genomes were assigned to global major lineages using pangolin v.2.0.5[15] to compare to the pangoLEARN database (version from 20th July 2020), default parameters were used for quality control filtering and probability scores for lineage calling.

## Results and Discussion

MinION sequencing generated complete genomic data sets for six of the eight sequenced isolates and for one swab out of two (Table 1). The remaining three samples (two isolates and one swab) demonstrated high Ct values when detecting viral cDNA by qPCR, suggesting that the concentrations of virus in these samples were low. PCR amplification was not uniform in these samples and genomic assemblies had short gaps where 30-fold coverage was not achieved. The primers used in the tiling protocol are unable to target the 3’ and 5’ extremities of the virus, and therefore all assemblies were missing 54 bases from the 5’ extremity and 67 bases from the 3’ extremity.

**Table 1.** Genome sequencing details.

| ID | Swab (S)<br>or Culture<br>(C) | Isolation/<br>Passage<br>Details<sup>+</sup> | MinION, amplicon tiling |  |  |  | Illumina |  |  |  | Combined |  |  |
| --- | --- | --- | --- | --- | --- | --- | --- | --- | --- | --- | --- | --- | --- |
|  |  |  | Complete | Missing | Ambiguities | Coverage** | Complete | Missing | Ambiguities | Coverage | Complete ? | Missing | Ambigui |
| hCoV-19/Réunion/RUN-PIMIT20 | C | Vero E6 | No | 1.7 % | 1 | 407x | Yes* | 2.1 % | 0 | 1500x | Yes* | 0.1 % | 0 |
| hCoV-19/Réunion/RUN-PIMIT1 | S | NA | Yes* | 0.4 % | 0 | 435x | Yes* | 2.1 % | 0 | 3250x | Yes* | 0.1 % | 0 |
| hCoV-19/Réunion/RUN-PIMIT2 | S | NA | No | 1.5 % | 1 | 356x | Yes* | 2.4 % | 0 | 675x | Yes* | 0.2 % | 0 |
| hCoV-19/Réunion/RUN-PIMIT8 | C | Vero E6 | Yes* | 0.4 % | 0 | 413x | Yes* | 0.1 % | 0 | 1700x | Yes* | 0.1 % | 0 |
| RUN-PIMIT1c | C | RUN-PIMIT1a,<br>passage 1 | Yes* | 0.4 % | 0 | 385x | - | - | - | - | - | - | - |
| RUN-PIMIT1d | C | RUN-PIMIT1c,<br>passage 2 | Yes* | 0.4 % | 0 | 423x | - | - | - | - | - | - | - |
| RUN-PIMIT1a | C | RUN-PIMIT1,<br>Vero E6 | No | 1.4 % | 0 | 336x | - | - | - | - | - | - | - |
| RUN-PIMIT1b | C | RUN-PIMIT1,<br>Vero E6 | Yes* | 0.4 % | 0 | 404x | - | - | - | - | - | - | - |
| RUN-PIMIT2a | C | RUN-PIMIT2,<br>Vero E6 | Yes* | 0.4 % | 0 | 394x | - | - | - | - | - | - | - |
| RUN-PIMIT20a | C | RUN-PIMIT20,<br>Vero E6 | Yes* | 0.4 % | 0 | 407x | - | - | - | - | - | - | - |

\* Genomes are regarded as complete if <2.5 % of the genome is missing from the 3' and 5' ends of the genome. Any genome containing consecutive ambiguities outside of the 3' and 5' regions is considered as incomplete.
\*\* MinION coverage is calculated by mapping in Medaka after subsampling sequence coverage to a maximum of 400x per primer pair. Overlapping regions increase average coverage.
\*\*\*Genomic sequences were considered as identical if no polymorphisms were present. Base calls in Illumina data versus ambiguous calls in MinION data are not considered to be contradictory (unless the ambiguity code is the base call in the Illumina sequence).
+ For cultured isolates, cell lines are indicated. For isolates that are resequenced from previous passages, corresponding parental strain is indicated.

Illumina shotgun sequencing produced near-complete genome assemblies for four of the samples (two isolates and two swabs), again only excluding 5’ and 3’ extremities of the genome, which had limited coverage that varied between isolates. By combining Illumina and MinION data mapped to the same reference strain we were able to improve the genomic assemblies for these four genome sequences. Furthermore, in genomic regions that were successfully sequenced by both approaches, no disagreements were observed in consensus sequences generated by MinION and Illumina sequencing. However, MinION assemblies had a slightly higher proportion of ambiguous base calls.

Sequencing data are summarized in Table 1.

Resequencing of isolated passage variants produced identical sequence variants to the parental strain in each instance (Table 2), suggesting that the process of viral isolation and passaging (in this case in Vero E6 cells) does not select for mutant variants.

**Table 2.**
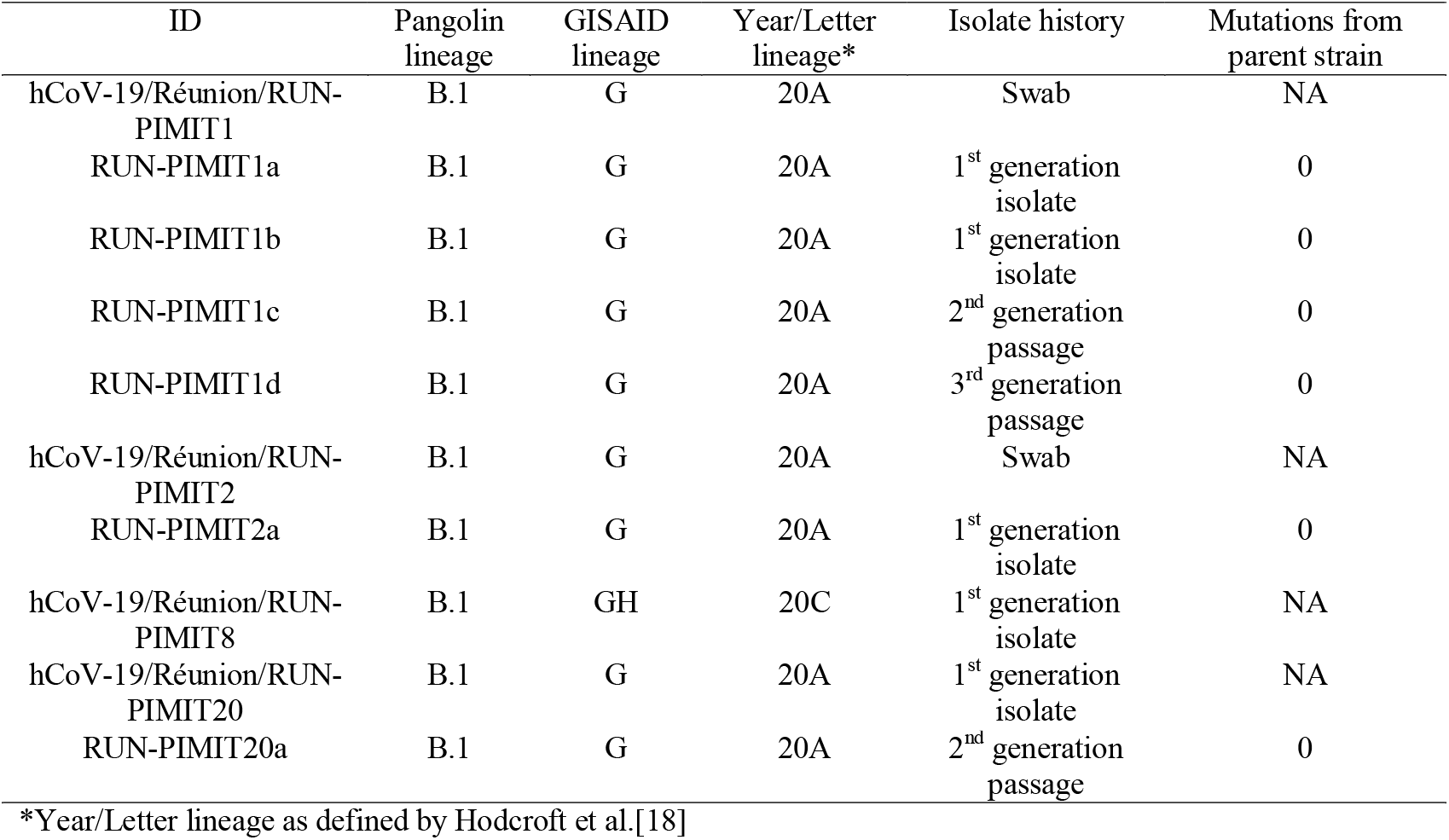
Lineage typing data of different isolates.

| ID | Pangolin lineage | GISAID lineage | Year/Letter lineage* | Isolate history | Mutations from parent strain |
| --- | --- | --- | --- | --- | --- |
| hCoV-19/Réunion/RUN-PIMIT1 | B.1 | G | 20A | Swab | NA |
| RUN-PIMIT1a | B.1 | G | 20A | 1<sup>st</sup> generation isolate | 0 |
| RUN-PIMIT1b | B.1 | G | 20A | 1<sup>st</sup> generation isolate | 0 |
| RUN-PIMIT1c | B.1 | G | 20A | 2<sup>nd</sup> generation passage | 0 |
| RUN-PIMIT1d | B.1 | G | 20A | 3<sup>rd</sup> generation passage | 0 |
| hCoV-19/Réunion/RUN-PIMIT2 | B.1 | G | 20A | Swab | NA |
| RUN-PIMIT2a | B.1 | G | 20A | 1<sup>st</sup> generation isolate | 0 |
| hCoV-19/Réunion/RUN-PIMIT8 | B.1 | GH | 20C | 1<sup>st</sup> generation isolate | NA |
| hCoV-19/Réunion/RUN-PIMIT20 | B.1 | G | 20A | 1<sup>st</sup> generation isolate | NA |
| RUN-PIMIT20a | B.1 | G | 20A | 2<sup>nd</sup> generation passage | 0 |

\*Year/Letter lineage as defined by Hodcroft et al.[18]

Typing assigned all genomes to pangolin lineage B.1, either belonging to GISAID lineage G, or GH (Table 2) which (along with lineage GR) are the prevailing lineages circulating globally at the time of writing [16]. Phylogenetic analysis identified three different sequence variants, which differed from each other by a maximum of seven mutations (Figure 2).

**Figure 2:**
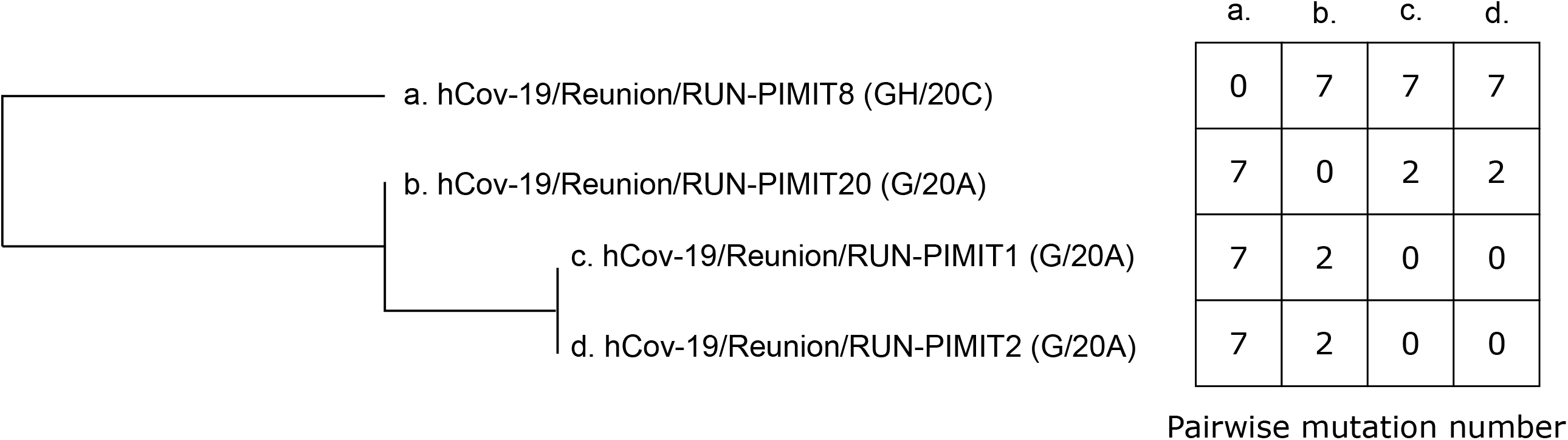
Phylogenetic tree and pairwise mutation number between genomes of SARS-CoV-2 from independent cases in La Reunion.

## Conclusion

Our findings provide support that MinION sequencing can be successfully applied for rapid sequencing and generation of genomic data in remote locations, such as on Reunion Island. Paired comparison with genomes obtained with Illumina sequencing further highlight that the quality of the generated sequences can be used confidently in molecular epidemiology and phylodynamic studies.

Additionally, we observed that viral isolation, limited passaging and production in Vero E6 cell lines were not associated with the selection of mutations that could bias genomic typing results. This is important due to the strong genetic similarity between prevailing SARS-CoV-2 lineages and because cell-line based viral amplification is a useful method for extracting increased quantities of viral material when direct sequencing of swabs with low viral loads may provide incomplete genomic data.

At the time of writing, Reunion Island is facing its first wave of COVID-19 infection, with more than 400 cases of COVID-19 reported weekly[17]. As has been demonstrated elsewhere, genomic data can be used to significantly improve the case assignment rate and contact tracing measures that help to mitigate future disease[6]. Phylodynamic investigations based on these data can also inform the timing and origin of viral introduction, as well as their local transmission dynamics and epidemiological characteristics. Thus, the capacity to perform in-house, rapid and comparatively cheap genomic typing of SARS-CoV-2 isolates will hopefully play an important role in improving Reunion Island’s response to the COVID-19 pandemic. These analyses are intended to continue on a larger local scale in Reunion Island in order to understand the circulation of SARS-CoV-2 during the current, largely autochthonous, COVID-19 epidemic wave.

## Data Availability

Genomic data from this work is available through the GISAID database.

## Acknowledgments

This research was funded by Université de La Réunion (COVID-19 emergency seed funding).

## Conflicts of interest

Authors declare no conflicts of interest.

## Notes

### Competing Interest Statement

The authors have declared no competing interest.

### Funding Statement

No external funding was received for this work.

### Author Declarations

Samples were obtained from the University Hospital Center of Reunion Island, after obtaining ethical committee approval (Comites de Protection des Personnes, Nord Ouest IV de Lille, France; number EudraCT / ID-RCB 2020-A01253-36). All participants gave their informed consent.

